# Regional Variation in the Interpretation of Contact Precautions for Multidrug-resistant Gram-negative bacteria: a cross-sectional survey

**DOI:** 10.1101/2024.03.29.24305013

**Authors:** Anneloes van Veen, Inge de Goeij, Marjolein Damen, Elisabeth G.W. Huijskens, Sunita Paltansing, Michiel van Rijn, Robbert G. Bentvelsen, Jacobien Veenemans, Michael van der Linden, Margreet C. Vos, Juliëtte A. Severin, the Infection Prevention and Antimicrobial Resistance Care Network South-western Netherlands

**Affiliations:** Department of Medical Microbiology and Infectious Diseases, Erasmus MC University Medical Centre, Rotterdam, The Netherlands; Department of Medical Microbiology, Maasstad General Hospital, Rotterdam, The Netherlands; Department of Medical Microbiology, Albert Schweitzer Hospital, Dordrecht, The Netherlands; Department of Medical Microbiology and Infection Prevention, Franciscus Gasthuis & Vlietland, Rotterdam, The Netherlands; Department of Medical Microbiology and Infectious Diseases, Ikazia Hospital, Rotterdam, The Netherlands; Department of Infection Prevention, Zorgsaam Hospital, Terneuzen, The Netherlands; Microvida Laboratory for Microbiology, Amphia Hospital, Breda, The Netherlands; Department of Infection Prevention, Admiraal de Ruyter Hospital, Goes, The Netherlands; Department of Infection Prevention, IJsselland Hospital, Capelle aan den IJssel, The Netherlands

**Keywords:** Carrier state, Enterobacteriaceae, Hospitals, Infection control, Multidrug resistance, Patient isolation, *Pseudomonas aeruginosa*

## Abstract

**Summary:** *Background:* Contact precautions (CP) are recommended when caring for patients with carbapenemase-producing Enterobacterales (CPE), carbapenemase-producing *Pseudomonas aeruginosa* (CPPA), and extended-spectrum β-lactamase-producing Enterobacterales (ESBL-E).

*Aim:* Our aim was to determine the interpretation of CP and associated infection prevention and control (IPC) measures in the non-ICU hospital setting for patients with CPE, CPPA or ESBL-E in 11 hospitals in the Southwest of the Netherlands.

*Methods:* A cross-sectional survey was developed to collect information on all implemented IPC measures, including use of personal protective equipment, IPC measures for visitors, cleaning and disinfection, precautions during outpatient care and follow-up strategies. All eleven hospitals were invited to participate between November 2020 and April 2021.

*Findings:* The survey was filled together with each hospital. All hospitals installed isolation precautions for patients with CPE and CPPA during inpatient care and day admissions, whereas ten hospitals (90.9%) applied isolation precautions for patients with ESBL-E. Gloves and gowns were always used during physical contact with the patient in isolation. Large variations were identified in IPC measures for visitors, cleaning and disinfection products used, and precautions during outpatient care. Four hospitals (36.4%) actively followed up on CPE or CPPA patients with the aim to declare them CPE- or CPPA-negative as timely as possible, and two hospitals (20.0%) actively followed up on ESBL-E patients.

*Conclusion:* CP are interpreted differently between hospitals, leading to regional differences in IPC measures applied in clinical settings. Harmonizing infection-control policies between the hospitals could facilitate patient transfers and benefit collective efforts of preventing transmission of MDR-GNB.

## Introduction

Multidrug-resistant Gram-negative bacteria (MDR-GNB), including carbapenemase-producing Enterobacterales (CPE), carbapenemase-producing *Pseudomonas aeruginosa* (CPPA), and extended-spectrum β-lactamase-producing Enterobacterales (ESBL-E), are well-known causes of healthcare-associated infections. Infections with MDR-GNB are more difficult to treat compared to infections with susceptible Gram-negative bacteria, and are, therefore, associated with high morbidity and mortality [1–3]. In the Netherlands, CPE and CPPA are rare, with only sporadic outbreaks in Dutch hospitals [4]. ESBL-E are more often found, but prevalences are still low in comparison to other European countries [4–6].

Infection prevention and control (IPC) measures are essential to prevent or control nosocomial spread of MDR-GNB. While there is in general international consensus on the application of contact precautions (CP), in addition to standard precautions, when caring for patients colonized or infected with MDR-GNB, international guidelines show some variation in their recommendations on related IPC measures, e.g., which personal protective equipment (PPE) healthcare workers (HCW) should use and when [7–14]. Also, IPC measures needed in specific settings (e.g., during outpatient care or physiotherapy of a hospitalized patient) and for different patient populations, are often not described, possibly due to a lack of studies in these specific situations and, consequently, paucity of evidence [11, 15, 16].

Besides variation in or lack of recommendations on certain IPC measures in infection-control guidelines, substantial variation in implemented IPC measures by healthcare facilities has been reported as well, both at the national and international level [11, 16–18]. Major differences were found, for example, in IPC measures between hospitals in a small geographical area [19]. The Infection Prevention and Antimicrobial Resistance (IP & AMR) Care Network South-western Netherlands is likewise a small geographical area, with a relatively large number of hospitals [20]. Patients are frequently transferred between these hospitals, which is a known risk factor for transmission of multidrug-resistant bacteria, and therefore necessitates a collaborative approach with consensus on IPC measures to prevent or control spread [15, 21–23]. The aim of this study was to determine the interpretation of CP and associated IPC measures in the non-ICU hospital setting for patients colonized or infected with CPE, CPPA or ESBL-E in the Southwest of the Netherlands.

## Methods

### Setting

This study was performed within the IP & AMR Care Network South-western Netherlands, which was established in 2015 as part of the Dutch AMR National Action Plan [24]. The eleven hospitals from the IP & AMR Care Network South-western Netherlands were invited to participate in this study, including one university hospital, six non-university teaching hospitals and four non-teaching hospitals (Supplementary Table A1).

### Survey

We developed a cross-sectional survey to collect information on the IPC measures embedded in the hospitals’ internal IPC policies for nine multidrug-resistant microorganisms (MRDO) [9]. Here, we only report on the IPC measures for CPE, CPPA, and ESBL-E. The survey focused on a variety of topics, including flagging of carriers in the electronic health record (EHR); isolation precautions during inpatient care, day admissions, and outpatient care; IPC measures for visitors of inpatients; terminal cleaning and disinfection; and follow-up of carriers and conditions for cessation of isolation measures. The majority of questions were multiple-choice, yet more detailed explanations could be given if necessary.

The survey was filled out together with one or multiple infection prevention practitioner(s) from each hospital during an online meeting between November 2020 and April 2021. In preparation of the meeting, the participants received the survey. After the meeting, the filled survey was sent to them by email to check whether the survey was filled out correctly. If necessary, answers could be added and modified after which the survey was sent back to the research team for analysis.

### Definitions

Supplementary Table A2 provides an overview of the IPC measures recommended by national and international IPC guidelines for CPE, CPPA, and ESBL-E. The Dutch guidelines describe different types of isolation, including strict isolation and contact isolation [25–27]. Currently, the Dutch MDRO guideline for hospitals indicates that inpatients with CPE, CPPA, and ESBL-E should be cared for in contact isolation, and specifically recommends using single-occupancy rooms when caring for patients with CPE [27]. Strict isolation is only recommended for multidrug-resistant *Acinetobacter* species. Furthermore, the Dutch MDRO guideline prescribes standard precautions, not targeted contact precautions, for patients visiting the outpatient clinic. Cessation of isolation measures is not recommended during hospitalization, although it could be considered if isolation is a major burden for the patient’s wellbeing and/or treatment. In this case, two negative culture sets, with at least 24 hours in between, are required [27].

For the follow-up of MDR-GNB carriers, active, passive and no follow-up are distinguished. Active follow-up is defined as requesting all MDR-GNB carriers to participate in taking screening cultures and actively following them with the aim to declare patients MDR-GNB-negative as timely as possible. During passive follow-up, however, patients only receive screening cultures on indication by the treating physician (e.g., when long-term isolation is detrimental to the patient’s health and/or treatment) in order to safely discontinue isolation measures following consecutive negative culture sets or are screened upon hospital admission. Hospitals not pursuing active or passive follow-up are categorized as no follow-up.

### Statistical analysis

IBM Statistical Package for the Social Sciences Solutions (SPSS) version 28 (IBM Corp., Armonk, New York, USA) was used for the analyses. Missing data are reported as such. For descriptive purposes, frequencies and percentages were calculated, where appropriate.

## Results

### Carbapenemase-producing Enterobacterales & carbapenemase-producing *Pseudomonas aeruginosa*

#### Inpatient care and day admissions

The survey was filled together with each of the 11 hospitals. All hospitals flagged patients colonized or infected with CPE and CPPA in their EHR. Infection-control policies for CPE and CPPA were in place in each hospital, however, the number and strength of IPC measures applied varied (Table I). During inpatient care and at day admissions, different types of isolation were applied: eight hospitals (72.7%) applied contact isolation, one hospital (9.1%) applied strict isolation, and two hospitals (18.2%) applied so-called “contact-plus” isolation (Table I). “Contact-plus” isolation is not defined in any national or international guideline. It is defined as isolation with measures that are in-between contact and strict isolation, as defined in the Dutch guidelines, but it is applied differently in two hospitals [25–27]. At hospital 8, the patient room’s door had to remain closed during “contact-plus” isolation, but could be open during contact isolation. “Contact-plus” isolation in hospital 1 differed from contact isolation in terms of the isolation measures required during outpatient care and the IPC measures for visitors. However, all hospitals that installed contact isolation for CPE and CPPA carriers also applied stricter measures than the Dutch contact isolation guideline prescribes (e.g., requiring HCWs to also wear a gown besides only gloves or requiring the patient room’s door to be closed), without defining it “contact-plus” isolation [26].

**Table I.** Transmission-based precautions during inpatient care and day admissions (n=11 hospitals).

| Transmission-based precautions | CPE | CPPA | ESBL-E |
| --- | --- | --- | --- |
| **<u>Isolation precautions during inpatient care</u>** |  |  |  |
| Yes | 11 (100) | 11 (100) | 10 (90.9) |
| No | 0 (0.0) | 0 (0.0) | 1 (9.1) |
| **Type of isolation** |  |  |  |
| Strict isolation | 1 (9.1) | 1 (9.1) | 0 (0.0) |
| Contact-plus isolation | 2 (18.2) | 2 (18.2) | 0 (0.0) |
| Contact isolation | 8 (72.7) | 8 (72.7) | 10 (90.9) |
| No isolation | 0 (0.0) | 0 (0.0) | 1 (9.1) |
| **<u>IPC measures during inpatient care</u>** |  |  |  |
| **Type of room<sup>1,2</sup>** |  |  |  |
| Isolation room with anteroom and private bathroom | 2 (18.2) | 2 (18.2) | 0 (0.0) |
| Single-patient room with private bathroom | 10 (90.9) | 10 (90.9) | 8 (72.7) |
| Single-patient room with shared bathroom | 1 (9.1) | 1 (9.1) | 1 (9.1) |
| Multiple-occupancy room with dedicated bathroom | 1 (9.1) | 1 (9.1) | 3 (27.3) |
| Multiple-occupancy room with shared bathroom | 0 (0.0) | 0 (0.0) | 3 (27.3) |
| Not applicable | 0 (0.0) | 0 (0.0) | 1 (9.1) |
| **HCWs PPE: entering the patient room without patient contact<sup>3</sup>** |  |  |  |
| No PPE | 5 (45.5) | 5 (45.5) | 8 (72.7) |
| Non-sterile gloves | 1 (9.1) | 1 (9.1) | 1 (9.1) |
| Disposable gown and non-sterile gloves | 4 (36.4) | 4 (36.4) | 1 (9.1) |
| Surgical mask type IIR, disposable gown, non-sterile gloves, and hair cap | 1 (9.1) | 1 (9.1) | 0 (0.0) |
| Not applicable | 0 (0.0) | 0 (0.0) | 1 (9.1) |
| **HCWs PPE: entering the patient room with patient contact<sup>3</sup>** |  |  |  |
| Gown and gloves | 10 (90.9) | 10 (90.9) | 10 (90.9) |
| IIR surgical mask, gown, gloves, and hair cap | 1 (9.1) | 1 (9.1) | 0 (0.0) |
| Not applicable | 0 (0.0) | 0 (0.0) | 1 (9.1) |
| **Room door allowed to be open** |  |  |  |
| Yes | 6 (54.5) | 7 (63.6) | 9 (81.8) |
| No | 5 (45.5) | 4 (36.4) | 1 (9.1) |
| Not applicable | 0 (0.0) | 0 (0.0) | 1 (9.1) |
| **Physiotherapy is allowed in public gym** |  |  |  |
| Yes | 5 (45.5) | 5 (45.5) | 7 (63.6) |
| No | 4 (36.4) | 4 (36.4) | 1 (9.1) |
| Not applicable | 2 (18.2) | 2 (18.2) | 3 (27.3) |
| **Physiotherapy in corridors and stairwells is allowed** |  |  |  |
| Yes | 8 (72.7) | 8 (72.7) | 10 (90.9) |
| No | 3 (27.3) | 3 (27.3) | 0 (0.0) |
| Not applicable | 0 (0.0) | 0 (0.0) | 1 (9.1) |
| Physiotherapist with PPE | 6 (75.0) | 6 (75.0) | 6 (60.0) |
| Patient with PPE | 1 (12.5) | 1 (12.5) | 1 (10.0) |
| No PPE for physiotherapist or patient | 1 (12.5) | 1 (12.5) | 2 (20.0)<sup>4</sup> |
| **Mandatory to report physiotherapy to infection prevention team** |  |  |  |
| Yes | 5 (55.6) | 5 (55.6) | 5 (45.5) |
| No | 4 (36.4) | 4 (36.4) | 5 (45.5) |
| Not applicable | 2 (18.2) | 2 (18.2) | 1 (9.1) |
| **<u>Isolation precautions during day admissions</u>** |  |  |  |
| Yes | 11 (100) | 11 (100) | 9 (81.8) |
| Only for a specific patient group | 0 (0.0) | 0 (0.0) | 1 (9.1) |
| No | 0 (0.0) | 0 (0.0) | 1 (9.1) |
| **Type of isolation** |  |  |  |
| Strict isolation | 1 (9.1) | 1 (9.1) | 0 (0.0) |
| Contact-plus isolation | 2 (18.2) | 2 (18.2) | 1 (9.1) |
| Contact isolation | 8 (72.7) | 8 (72.7) | 9 (81.8) |
| No isolation | 0 (0.0) | 0 (0.0) | 1 (9.1) |
| **<u>IPC measures during day admissions</u>** |  |  |  |
| **Type of room<sup>5</sup>** |  |  |  |
| Single-patient room | 9 (81.8) | 9 (81.8) | 5 (45.5) |
| Multiple-occupancy room | 1 (9.1) | 1 (9.1) | 4 (36.4) |
| Not applicable | 1 (9.1) | 1 (9.1) | 2 (18.2) |
| **HCW PPE in treatment room<sup>3</sup>** |  |  |  |
| No PPE | 0 (0.0) | 0 (0.0) | 0 (0.0) |
| Non-sterile gloves | 0 (0.0) | 0 (0.0) | 0 (0.0) |
| Disposable gown and non-sterile gloves | 10 (90.9) | 10 (90.9) | 10 (90.9) |
| Surgical mask type IIR, disposable gown, non-sterile gloves, and hair cap | 1 (9.1) | 1 (9.1) | 0 (0.0) |
| Not applicable | 0 (0.0) | 0 (0.0) | 1 (9.1) |
| **Allowed to be accompanied by a visitor** |  |  |  |
| Yes | 11 (100) | 11 (100) | 10 (90.9) |
| No | 0 (0.0) | 0 (0.0) | 0 (0.0) |
| Not applicable | 0 (0.0) | 0 (0.0) | 1 (9.1) |
| **Visitor also placed in isolation** |  |  |  |
| Yes | 6 (54.5) | 6 (54.5) | 4 (36.4) |
| No | 5 (45.5) | 5 (45.5) | 6 (54.5) |
| Not applicable | 0 (0.0) | 0 (0.0) | 1 (9.1) |

Data are N (%). CPPA carbapenemase-producing *Pseudomonas aeruginosa*, CPE carbapenemase-producing Enterobacterales, ESBL-E extended-spectrum β-lactamase-producing Enterobacterales, HCW healthcare worker, IPC infection prevention and control, PPE personal protective equipment.
<sup>1</sup> Multiple answers per hospital possible.
<sup>2</sup> An isolation room is defined as a single-occupancy room with anteroom and pressure difference. A multiple-occupancy patient room is considered to be a room in which other bed(s) may be occupied both by carriers of the same or different resistant bacteria, and non-carriers.
<sup>3</sup> A disposable gown was defined as a long-sleeved gown in 10 out of 11 hospitals. Hospital 9 used a long-sleeved gown when caring for patients in strict isolation, which this hospital applied to patients with CPE or CPPA, and an apron when caring for patients in contact isolation, which this hospital applied to patients with ESBL-E.
<sup>4</sup> One answer is missing.
<sup>5</sup> Procedures taking place in the day admissions centre are e.g., minor surgical procedures, haemodialysis, hemapheresis, etc. The answer is not applicable when a patient directly goes home after the procedure has been performed in an operating room.

Most hospitals isolated CPE and CPPA patients during inpatient care in single-occupancy rooms with private bathrooms (n=10, 90.9%). While according to the Dutch guidelines an isolation room is only necessary for strict isolation, one hospital (9.1%) reported isolating CPE and CPPA carriers in isolation rooms, but still used the term contact isolation (Table I) [25]. Patients were preferably isolated in single-occupancy rooms during day admissions (n=9, 81.8%), although shortage of such rooms could necessitate isolation in multiple-occupancy rooms (n=1, 9.1%).

Although in five hospitals (45.5%) the decision for HCW to put on PPE before entering a patient’s room depended on whether contact with the patient was anticipated or not, in other hospitals HCW were required to wear PPE at all times when entering the patient’s room (n=6, 54.5%). Overall, ten hospitals (90.9%) were more stringent compared to the Dutch contact isolation guideline, which requires HCW to wear only gloves before having contact with the patient and/or the patient’s environment [26].

#### Outpatient care

Four hospitals (36.4%) only used standard precautions when patients with CPE or CPPA visited the outpatient clinic. Hospital 1 (9.1%) consistently applied “contact-plus” isolation during outpatient care (policy 1), while the other six hospitals (54.5%) followed different approaches to determine the type of isolation (policy 2-5; Figure 1). These decisions depended on the type of patient visiting the outpatient clinic, the type of procedure being performed, and/or on whether there would be physical contact with the patient. In general, contact isolation was applied during invasive procedures and inpatients were continued to be cared for in contact isolation when visiting the outpatient clinic. During contact isolation, HCW were wearing a disposable gown and non-sterile gloves (Figure 1).

**Figure 1.**
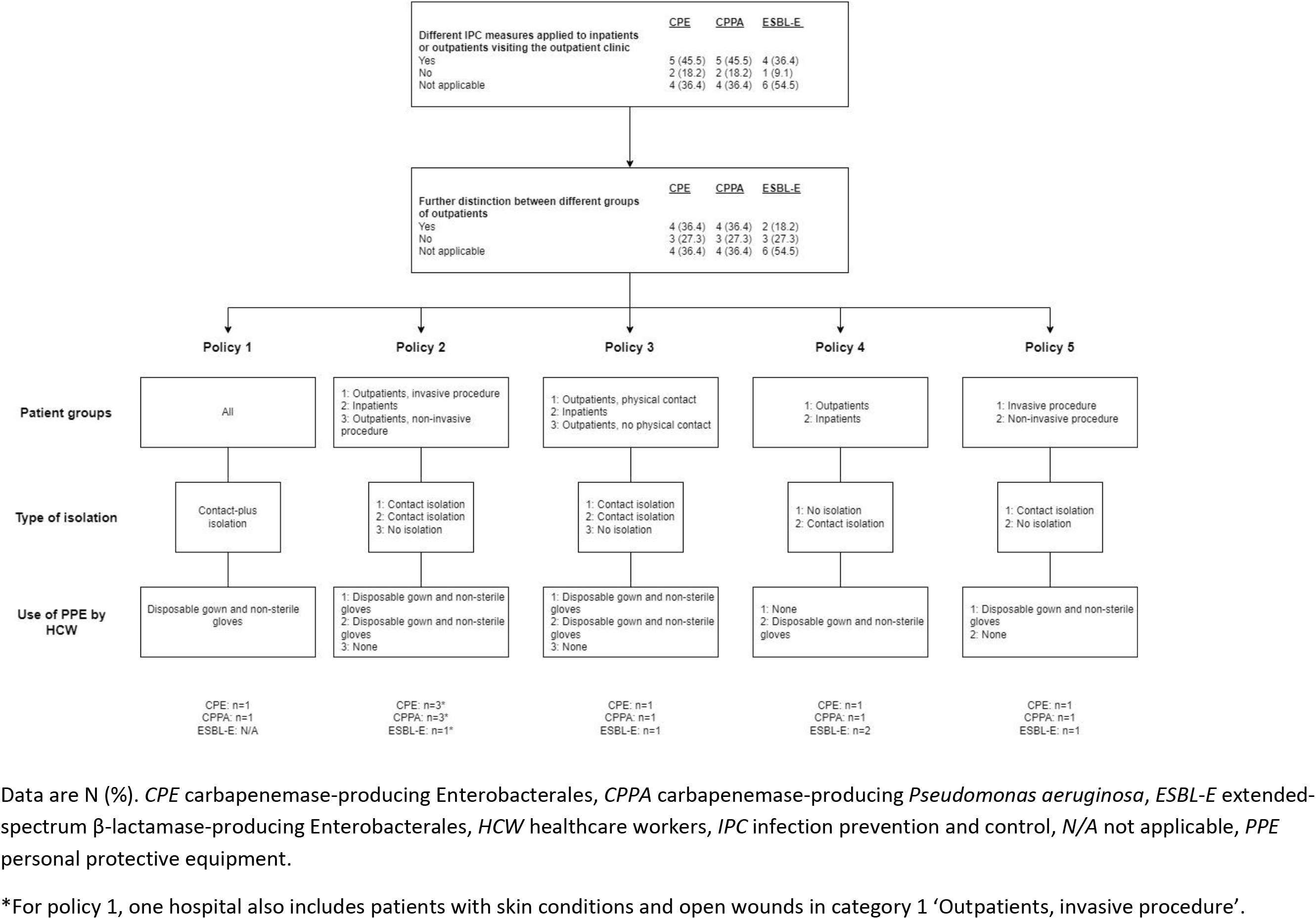
Contact precautions during outpatient care (n=11 hospitals). Data are N (%). *CPE* carbapenemase-producing Enterobacterales, *CPPA* carbapenemase-producing *Pseudomonas aeruginosa*, *ESBL-E* extended-spectrum β-lactamase-producing Enterobacterales, *HCW* healthcare workers, *IPC* infection prevention and control, *N/A* not applicable, *PPE* personal protective equipment. *For policy 1, one hospital also includes patients with skin conditions and open wounds in category 1 ‘Outpatients, invasive procedure’.

#### IPC measures for visitors

Visitors were requested to take precautions when visiting an inpatient with CPE or CPPA in all hospitals, ranging from hand disinfection (n=11, 100%) to, additionally, wearing a surgical mask, disposable gown, non-sterile gloves and hair cap as the most extensive IPC measures (n=1, 9.1%). In four hospitals (36.4%), visitors could temporarily leave a patient’s room during their visit. Rooming-in was permitted in most hospitals (n=10, 90.9%), of which five hospitals (50.0%) allowed rooming-in visitors to temporarily leave the patient’s room (Supplementary Table A3).

#### Cleaning & disinfection

Different cleaning and disinfection products were used (Supplementary Table A4). Most hospitals (n=9, 81.8%) replaced the separation curtains after cessation of isolation measures. One hospital (9.1%) also changed the window curtains (Supplementary Table A4).

#### Follow-up of carriage and conditions for cessation of isolation measures

Various follow-up strategies were used (Table II). Active follow-up was done by four hospitals (36.4%), wherein patients received a self-sampling set at home at different time intervals. Three hospitals (75.0%) initiated active follow-up two months after a positive culture, yet one hospital (25.0%) started at least one year after the last positive culture. The number of swabs needed to declare a patient CPE- or CPPA-negative varied between these four hospitals, from one negative culture to six consecutive negative culture sets. Passive follow-up was performed in another four hospitals (36.4%). These hospitals all used two consecutive negative cultures or culture sets for lifting isolation measures, but the initiation of follow-up varied from at least 48 hours after stopping relevant antibiotics to at least one year after the last positive culture. Although negative cultures could lead to declaring a patient CPE- or CPPA-negative followed by cessation of isolation measures, two hospitals (50.0%) continued to flag patient’s EHR indicating former CPE- or CPPA-carriage. Three hospitals (27.3%) did not follow up on CPE or CPPA carriers and flagged patient’s EHR indefinitely (Table II).

**Table II.** Follow-up and conditions for cessation of isolation measures for patients with CPE and CPPA (n=11 hospitals).

|  | Hospital<br>1 | Hospital<br>4 | Hospital<br>5 | Hospital<br>10 | Hospital<br>2 | Hospital<br>3 | Hospital<br>6 | Hospital<br>11 | Hospital<br>7 | Hospital<br>8 | Hospital<br>9 |
| --- | --- | --- | --- | --- | --- | --- | --- | --- | --- | --- | --- |
| Contraindications for initiation of active follow-up or culturing | Yes, antibiotic use and hospital admission | Yes, antibiotic use | None | Yes, antibiotic use | Yes, antibiotic use and hospital admission | Yes, antibiotic use; hospital admission only for CPPA | Yes, antibiotic use | Yes, antibiotic use, drains, and wounds | N/A | N/A | N/A |
| Active, passive or no follow-up | Active | Active | Active | Active | Passive | Passive | Passive | Passive | No follow-up | No follow-up | No follow-up |
| Method of approaching patients | Self-sampling set by regular mail | Letter, followed by self-sampling set by regular mail | Self-sampling set by regular mail | Self-sampling set by regular mail | N/A | N/A | N/A | N/A | N/A | N/A | N/A |
| Number of follow-up swabs | 6 sets of 2 swabs | 5 sets of 1 swab | 1 set of 2 swabs | 5 sets of 1 swab | 2 sets of 3 swabs | 2 sets of 1 swab | 2 sets of 1 swab | 2 sets of 1 swab | N/A | N/A | N/A |
| Body sites<br>sampled | Throat,<br>rectum,<br>wounds (if<br>present),<br>urine and<br>sputum for<br>CPE (if<br>applicable) | Rectum,<br>urine only<br>in case of<br>UTI or<br>catheter | Throat<br>and<br>rectum | Rectum<br>(or<br>faeces) | Throat,<br>nose,<br>rectum,<br>wounds (if<br>present),<br>urine (if<br>catheter),<br>sputum (if<br>productive<br>cough,<br>intubation<br>or<br>tracheo-<br>stomy) | Rectum | Rectum | Rectum;<br>wounds<br>(if<br>present)<br>for CPE | N/A | N/A | N/A |
| Start active follow-up or culturing | Two months<br>after first<br>positive<br>culture and<br>without<br>contra-<br>indications | Two<br>months<br>after the<br>last<br>positive<br>culture | At least<br>one year<br>after the<br>last<br>positive<br>culture | Two<br>months<br>after the<br>last<br>positive<br>culture | 48 hours<br>after<br>stopping<br>antibiotics | Two<br>months<br>after the<br>last<br>positive<br>culture | At least 48<br>hours after<br>stopping<br>antibiotics | At least<br>one year<br>after the<br>last<br>positive<br>culture | N/A | N/A | N/A |
| Intervals | Every two months | Every two months | N/A | Every two months | At least 24 hours | Two days | 24 hours | At least 24 hours | N/A | N/A | N/A |
| Unflagging patient's EHR | After 6 negative culture sets | After 5 negative culture sets within 1 year | After 1 year with 1 negative culture set | After 5 negative cultures | After 2 negative culture sets<sup>1</sup> | After 2 negative cultures | Never<sup>2</sup> | After 2 negative culture sets | Never | Never | Never |

*CPPA* carbapenemase-producing *Pseudomonas aeruginosa*, CPE carbapenemase-producing Enterobacterales, N/A not applicable, UTI urinary tract infection.
Blue columns indicate passive follow-up of carriers. Number and frequency of cultures described in the blue columns represents the cultures needed for safe cessation of isolation measures during hospitalization.
<sup>1</sup> Electronic label is changed to 'Multidrug-resistant microorganism in the past', which means that patients are screened during each subsequent hospitalization.
<sup>2</sup> Electronic label remains at all times, however, isolation measures are scaled down in case screening cultures are negative after one year. Patients are screened during each subsequent hospitalization.

### Extended-spectrum β-lactamase-producing Enterobacterales

#### Inpatient care and day admissions

Patients with ESBL-E were also flagged in each hospital’s EHR. Hospital 2 (9.1%) did not apply isolation precautions for ESBL-E carriers (Table I). Hospital 1 (9.1%) placed all patient groups with ESBL-E during inpatient care in contact isolation, whereas isolation precautions varied for different patient groups during day admissions. Inpatients undergoing a procedure at the day admissions centre remained in contact isolation, whereas patients coming from home were cared for taking only standard precautions. Hospital 10 (9.1%) installed contact isolation for ESBL-E patients during inpatient care, yet “contact-plus” isolation during day admissions. Lastly, eight hospitals (72.7%) applied contact isolation for all patients with ESBL-E during both inpatient care and day admissions. Although a single-occupancy room with private bathroom remained preferable for inpatients with ESBL-E (n=8, 72.7%), several hospitals (n=6, 54.5%) also allowed these patients to stay in a multiple-occupancy room with or without dedicated bathroom due to shortage of single-occupancy rooms (Table I).

The policy of PPE use by HCW was more constant over the hospitals. Approximately 70% of hospitals (n=8) did not require HCW to wear any PPE upon room entrance when no patient contact was anticipated, while ten hospitals (90.9%) required HCW to wear a gown and gloves when contact was anticipated (Table I).

#### Outpatient care

Six hospitals (54.5%) did not take isolation precautions for ESBL-E carriers during outpatient care (Figure 1). Hospital 1 and 11 (18.2%) only placed inpatients with an appointment at the outpatient department in contact isolation, whereas outpatients were cared for using only standard precautions (policy 4). The other three hospitals (27.3%) followed a similar two- or three category approach as used for patients with CPE and CPPA (policy 2, 3, and 5; Figure 1).

#### IPC measures for visitors

Ten hospitals (90.9%) requested visitors to apply IPC measures, of which all hospitals imposed hand disinfection. Hospital 10 (10.0%) asked visitors to additionally wear a gown and gloves. Rooming-in was allowed in most hospitals (n=9, 81.8%). Whether temporary room leave by visitors was allowed, varied depending on the underlying reason and whether it was a regular or rooming-in visitor (Supplementary Table A3).

#### Cleaning & disinfection

A variety of cleaning and disinfection products was used after cessation of isolation measures (Supplementary Table A4). The separation curtains were replaced in seven hospitals (63.6%), while none of the hospitals changed the window curtains after cessation of isolation measures (Supplementary Table A4).

#### Follow-up of carriage and conditions for cessation of isolation measures

Four different strategies were used for follow-up of ESBL-E carriers (Table III). Two hospitals (18.2%) performed an active follow-up with varying duration and number of culture sets required. Whereas hospital 1 required two negative culture sets starting two months after the first positive culture with three days between culture sets, hospital 5 required one negative culture set at least one year after the last positive culture. Passive follow-up of ESBL-E carriers was performed by six hospitals (54.5%), with three hospitals (50.0%) screening on indication by the treating physician and three hospitals (50.0%) upon hospital admission. The former three hospitals all required two negative culture sets for the safe cessation of isolation measures, although the timing of when to start culturing varied. Two hospitals (18.2%) did not pursue any follow-up and unflagged the EHR automatically after one year.

**Table III.**
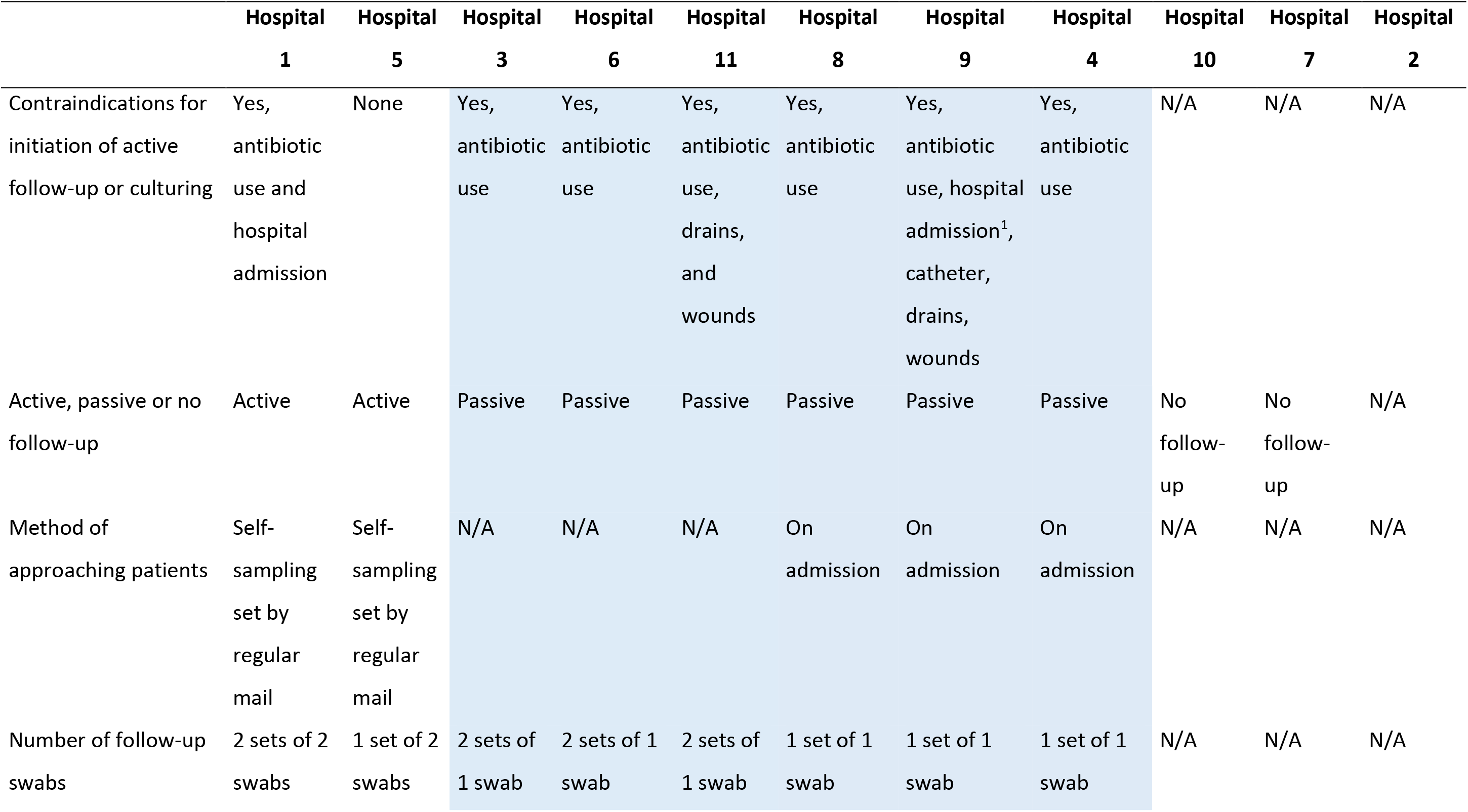
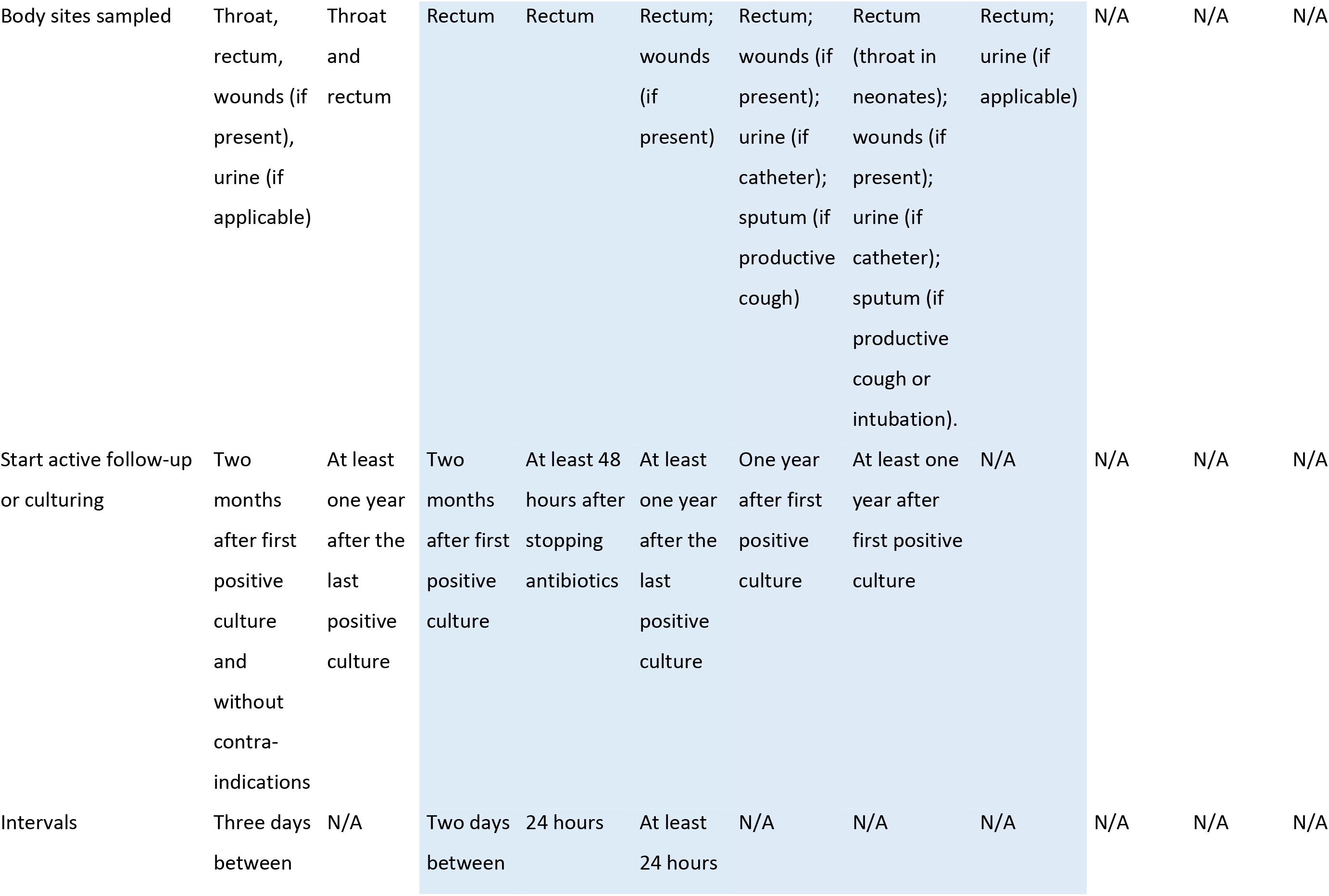
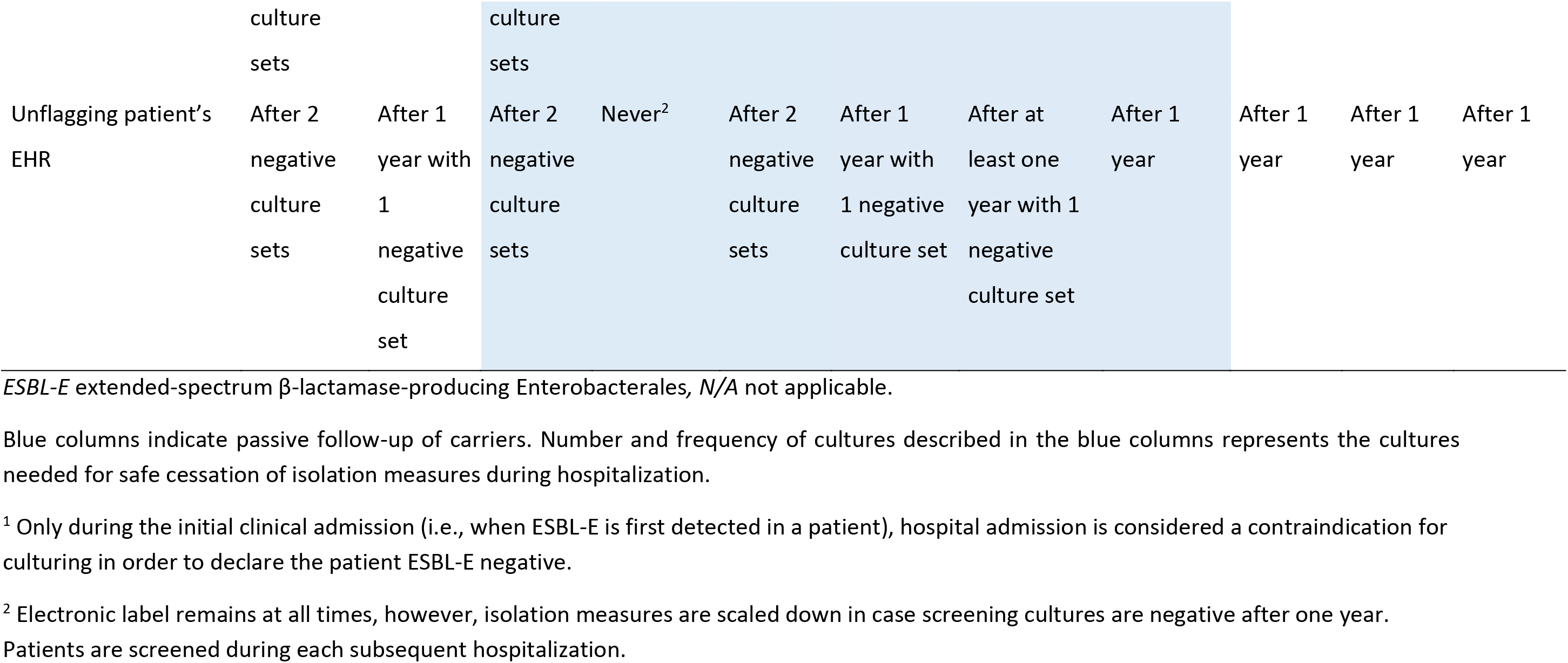
Follow-up and conditions for cessation of isolation measures for patients with ESBL-E (n=11 hospitals).

## Discussion

This study showed substantial variation in the interpretation of CP and associated IPC measures for patients with CPE, CPPA and ESBL-E in the non-ICU setting in hospitals in the Southwest of the Netherlands. Unsurprisingly, most variation was observed in the IPC measures applied in clinical settings, which are not well-defined in national and/or international guidelines.

The hospitals had different interpretations of CP, which is particularly highlighted by the introduction of a new type of isolation, “contact-plus” isolation, and the variation observed in associated IPC measures outlined in each hospital’s internal IPC policies. Similar to previous findings, most and most stringent IPC measures are applied for patients with CPE and CPPA compared to ESBL-E [11, 18, 19]. Variability in the type of room used, types of PPE used by HCW, and environmental cleaning regimens were demonstrated between hospitals for each MDR-GNB, which also confirms findings from previous studies [11, 16–19]. Moreover, considerable variation was observed in the IPC measures taken during outpatient care, with some hospitals installing no isolation precautions and other hospitals following a stepwise approach to determine which type of isolation, if any, was required for each patient.

Evidence-based recommendations on follow-up of MDR-GNB carriers and conditions for safe cessation of isolation measures are unavailable in international infection-control guidelines, possibly due to scarcity of data on the duration of colonization and the occurrence of relapse of recolonization. The Dutch MDRO guideline also offers limited guidance [27]. This is reflected in the large variety of follow-up strategies within our region, which varied from actively reaching out to patients with the aim to declare patients MDR-GNB-negative as timely as possible to no follow-up and flagging the EHR indefinitely. [28–31]. A direct comparison between different follow-up strategies in terms of duration and number of culture sets could be of added value in efforts to harmonize follow-up strategies.

In general, variation in implemented IPC measures on international, national, and regional levels, seems to depend on the availability of IPC guidelines for MDR-GNB, the evidence-base and level of detail of the IPC measures recommended in these guidelines, local context and epidemiology, and organizational resources (e.g., number and availability of single-occupancy rooms and personnel) [16–19]. National and international guidelines are available, yet, in practice, individual hospitals seem to apply the recommended measures quite differently. For example, hospital 2 does not apply CP for ESBL-E carriers, which is not in line with both the Dutch and European Society of Clinical Microbiology and Infectious Diseases (ESCMID) guidelines (with the exception of *Escherichia coli)*, and the active follow-up performed by several hospitals is not described in any national or international guideline whatsoever. Some regional differences can be explained by the patient populations served by the hospitals. Hospital 1 and 4, for example, provide tertiary care for critically-ill patients, resulting in a more intensive follow-up to prevent nosocomial transmissions among their vulnerable patients. The smaller hospitals may have less resources available and are, therefore, restricted in pursuing such intensive efforts. Also, hospital 1 and 4 have 100% adult single-occupancy rooms available, whereas the other, especially smaller, hospitals may need to deviate from their own and other hospitals’ policies, by occasionally placing MDR-GNB-positive patients in multiple-occupancy rooms due to shortage of single-occupancy rooms.

The observed regional variety shows the need to harmonize IPC measures, since one hospital’s actions (or non-actions) may impact other hospitals in the region that share patients [32]. Harmonizing measures, including conditions for cessation of isolation measures, may have a positive effect on MDR-GNB prevalence, may facilitate inter-hospital communication, provides HCWs clarity during patient transfers, and can lead to quicker and more effective actions to prevent or stop MDR-GNB spread between hospitals [19, 33, 34]. Also, it may cause more understanding and acceptance of IPC measures by patients (and their visitors) when they receive care in different hospitals, which may cause higher compliance with the installed measures. Research has shown that enhanced coordination in infection prevention within a region leads to greater synergistic effects, benefiting both individual hospitals and the entire region [32, 35–37].

However, which particular IPC measures are most effective and should thus be prioritised in specific settings remains somewhat unclear. Studies on the effectiveness of CP often lack information on the details of CP applied in specific settings, complicating the interpretation of their results [38, 39]. For example, whether a dedicated bathroom was used and whether terminal cleaning and disinfection of that bathroom was performed after cessation of isolation measures is frequently not described, while its importance is increasingly recognized. Therefore, authors should provide more details about which specific IPC measures are considered part of CP in their studies.

### Strengths and limitations

A strength of this study is its multicentre design, with all hospitals in our region participating. This allowed for a comprehensive overview of the IPC measures applied in the hospitals. Additionally, the extensiveness of the survey enabled us to uncover and compare details of IPC components associated with CP and isolation. A limitation of this study is that we did not ask the hospitals about the underlying reasons for choices made regarding their IPC policies. Future efforts should be aimed at harmonizing regional IPC policies, followed by measurement of each hospital’s compliance with these policies.

## Conclusions

Hospitals in the Southwest of the Netherlands reported considerable variation in the interpretation of CP and associated IPC measures when caring for patients with CPE, CPPA and ESBL-E. Heterogeneity in policies appeared most prevalent for isolation precautions during outpatient care and follow-up of carriers, which are not (well-)defined in national and/or international guidelines. Future research should explore the setting-specific reasons and risks related to these differences in IPC measures. Harmonizing infection-control policies between hospitals could facilitate patient transfers and benefit collective efforts of preventing MDR-GNB transmission.

## Data Availability

All data produced in the present study are available upon reasonable request to the authors.

## Acknowledgements

The authors would like to acknowledge the contributions of Janet Vos, former network manager of the IP & AMR Care Network South-western Netherlands, and all infection prevention practitioners from participating hospitals.

## Supplementary material

Supplementary Table A1: Characteristics of the eleven hospitals from the Southwest region of the Netherlands.

Supplementary Table A2: IPC measures recommended by the Dutch, ESCMID and ECDC guidelines.

Supplementary Table A3: IPC measures for visitors of inpatients (n=11 hospitals).

Supplementary Table A4: Cleaning and disinfection after discharge of the patient (n=11 hospitals).

## References

[1] Antimicrobial Resistance C. Global burden of bacterial antimicrobial resistance in 2019: a systematic analysis. Lancet 2022;399(10325):629–55.

[2] Borer A, Saidel-Odes L, Riesenberg K, Eskira S, Peled N, Nativ R, et al. Attributable mortality rate for carbapenem-resistant *Klebsiella pneumoniae* bacteremia. Infect Control Hosp Epidemiol 2009;30(10):972–6.

[3] Saharman YR, Pelegrin AC, Karuniawati A, Sedono R, Aditianingsih D, Goessens WHF, et al. Epidemiology and characterisation of carbapenem-non-susceptible *Pseudomonas aeruginosa* in a large intensive care unit in Jakarta, Indonesia. Int J Antimicrob Agents 2019;54(5):655–60.

[4] Nethmap/MARAN. NethMap 2021: Consumption of antimicrobial agents and antimicrobial resistance among medically important bacteria in the Netherlands. 2021.

[5] European Centre for Disease Prevention and Control. European Antimicrobial Resistance Surveillance Network (EARS-Net), https://www.ecdc.europa.eu/en/about-us/networks/disease-networks-and-laboratory-networks/ears-net-data; 2021 [accessed April 4 2023.].

[6] van der Schoor AS, Severin JA, Klaassen CHW, van den Akker JPC, Bruno MJ, Hendriks JM, et al. Universal screening or a universal risk assessment combined with risk-based screening for multidrug-resistant microorganisms upon admission: Comparing strategies. PLoS One 2023;18(7):e0289163.

[7] Tacconelli E, Cataldo MA, Dancer SJ, De Angelis G, Falcone M, Frank U, et al. ESCMID guidelines for the management of the infection control measures to reduce transmission of multidrug-resistant Gram-negative bacteria in hospitalized patients. Clin Microbiol Infect 2014;20 Suppl 1:1–55.

[8] Siegel JD, Rhinehart E, Jackson M, Chiarello L, Health Care Infection Control Practices Advisory C. 2007 Guideline for Isolation Precautions: Preventing Transmission of Infectious Agents in Health Care Settings. Am J Infect Control 2007;35(10 Suppl 2):S65–164.

[9] Kluytmans-Vandenbergh MF, Kluytmans JA, Voss A. Dutch guideline for preventing nosocomial transmission of highly resistant microorganisms (HRMO). Infection 2005;33(5-6):309–13.

[10] Magiorakos AP, Burns K, Rodríguez Baño J, Borg M, Daikos G, Dumpis U, et al. Infection prevention and control measures and tools for the prevention of entry of carbapenem-resistant Enterobacteriaceae into healthcare settings: guidance from the European Centre for Disease Prevention and Control. Antimicrob Resist Infect Control 2017;6:113.

[11] Lynch BL, Schaffer K. Can guidelines for the control of multi-drug-resistant Gram-negative organisms be put into practice? A national survey of guideline compliance and comparison of available guidelines. J Hosp Infect 2019;102(1):1–7.

[12] HSE Antimicrobial Resistance and Infection Control. Management and Control of Carbapenemase Producing Enterobacterales (CPE) in all Healthcare Settings. Dublin: HSE-AMRIC;; December 2022.

[13] Otter JA, Mutters NT, Tacconelli E, Gikas A, Holmes AH. Controversies in guidelines for the control of multidrug-resistant Gram-negative bacteria in EU countries. Clin Microbiol Infect 2015;21(12):1057–66.

[14] Centers for Disease Control. Facility guidance for control of carbapenem-resistant Enterobacteriaceae (CRE): November 2015 update - CRE Toolkit. 2015.

[15] European Centre for Disease Prevention and Control. Risk assessment on the spread of carbapenemase-producing Enterobacteriaceae (CPE) through patient transfer between healthcare facilities, with special emphasis on cross-border transfer. 2011.

[16] Tacconelli E, Buhl M, Humphreys H, Malek V, Presterl E, Rodriguez-Baño J, et al. Analysis of the challenges in implementing guidelines to prevent the spread of multidrug-resistant gram-negatives in Europe. BMJ Open 2019;9(5):e027683.

[17] van Dijk MD, Voor In ’t Holt AF, Alp E, Hell M, Petrosillo N, Presterl E, et al. Infection prevention and control policies in hospitals and prevalence of highly resistant microorganisms: an international comparative study. Antimicrob Resist Infect Control 2022;11(1):152.

[18] Gysin DV, Cookson B, Saenz H, Dettenkofer M, Widmer AF, Infections ESGfN. Variability in contact precautions to control the nosocomial spread of multi-drug resistant organisms in the endemic setting: a multinational cross-sectional survey. Antimicrob Resist Infect Control 2018;7:81.

[19] Tschudin-Sutter S, Lavigne T, Grundmann H, Rauch J, Eichel VM, Deboscker S, et al. Differences in infection control and diagnostic measures for multidrug-resistant organisms in the tristate area of France, Germany and Switzerland in 2019 - survey results from the RH(E)IN-CARE network. Swiss Med Wkly 2021;151:w20454.

[20] Infectiepreventie & Antibioticaresistentie Zorgnetwerk Zuidwest-Nederland. Aanpak antibioticaresistentie en infectiepreventie Zuidwest-Nederland, https://abrzorgnetwerkzwn.nl/; [accessed Feb 13 2024].

[21] Rogers BA, Aminzadeh Z, Hayashi Y, Paterson DL. Country-to-country transfer of patients and the risk of multi-resistant bacterial infection. Clin Infect Dis 2011;53(1):49–56.

[22] Donker T, Wallinga J, Grundmann H. Patient referral patterns and the spread of hospital-acquired infections through national health care networks. PLoS Comput Biol 2010;6(3):e1000715.

[23] Ciccolini M, Donker T, Köck R, Mielke M, Hendrix R, Jurke A, et al. Infection prevention in a connected world: the case for a regional approach. Int J Med Microbiol 2013;303(6-7):380–7.

[24] Tweede Kamer der Staten-Generaal. Kamerbrief 32620, nr. 159. 2015.

[25] Dutch Working Party on Infection Prevention (WIP). Strikte isolatie. 2006.

[26] Dutch Working Party on Infection Prevention (WIP). Contactisolatie. 2006.

[27] Dutch Working Party on Infection Prevention (WIP). Bijzonder resistente micro-organismen (BRMO). 2012.

[28] van Dijk MD, Voor In ’t Holt AF, Polinder S, Severin JA, Vos MC. The daily direct costs of isolating patients identified with highly resistant micro-organisms in a non-outbreak setting. J Hosp Infect 2021;109:88–95.

[29] Roth JA, Hornung-Winter C, Radicke I, Hug BL, Biedert M, Abshagen C, et al. Direct Costs of a Contact Isolation Day: A Prospective Cost Analysis at a Swiss University Hospital. Infect Control Hosp Epidemiol 2018;39(1):101–3.

[30] Mehrotra P, Croft L, Day HR, Perencevich EN, Pineles L, Harris AD, et al. Effects of contact precautions on patient perception of care and satisfaction: a prospective cohort study. Infect Control Hosp Epidemiol 2013;34(10):1087–93.

[31] Stelfox HT, Bates DW, Redelmeier DA. Safety of patients isolated for infection control. Jama 2003;290(14):1899–905.

[32] Lee BY, Yilmaz SL, Wong KF, Bartsch SM, Eubank S, Song Y, et al. Modeling the regional spread and control of vancomycin-resistant enterococci. Am J Infect Control 2013;41(8):668–73.

[33] Jurke A, Daniels-Haardt I, Silvis W, Berends MS, Glasner C, Becker K, et al. Changing epidemiology of meticillin-resistant *Staphylococcus aureus* in 42 hospitals in the Dutch-German border region, 2012 to 2016: results of the search-and-follow-policy. Euro Surveill 2019;24(15).

[34] Jurke A, Kock R, Becker K, Thole S, Hendrix R, Rossen J, et al. Reduction of the nosocomial meticillin-resistant *Staphylococcus aureus* incidence density by a region-wide search and follow-strategy in forty German hospitals of the EUREGIO, 2009 to 2011. Euro Surveill 2013;18(36):pii=20579.

[35] Bartsch SM, Wong KF, Mueller LE, Gussin GM, McKinnell JA, Tjoa T, et al. Modeling Interventions to Reduce the Spread of Multidrug-Resistant Organisms Between Health Care Facilities in a Region. JAMA Netw Open 2021;4(8):e2119212.

[36] Lee BY, Bartsch SM, Wong KF, McKinnell JA, Slayton RB, Miller LG, et al. The Potential Trajectory of Carbapenem-Resistant Enterobacteriaceae, an Emerging Threat to Health-Care Facilities, and the Impact of the Centers for Disease Control and Prevention Toolkit. Am J Epidemiol 2016;183(5):471–9.

[37] Lee BY, Bartsch SM, Wong KF, Yilmaz SL, Avery TR, Singh A, et al. Simulation shows hospitals that cooperate on infection control obtain better results than hospitals acting alone. Health Aff (Millwood) 2012;31(10):2295–303.

[38] Maechler F, Schwab F, Hansen S, Fankhauser C, Harbarth S, Huttner BD, et al. Contact isolation versus standard precautions to decrease acquisition of extended-spectrum β-lactamase-producing Enterobacterales in non-critical care wards: a cluster-randomised crossover trial. Lancet Infect Dis 2020;20(5):575–84.

[39] Kluytmans-van den Bergh MFQ, Bruijning-Verhagen PCJ, Vandenbroucke-Grauls C, de Brauwer E, Buiting AGM, Diederen BM, et al. Contact precautions in single-bed or multiple-bed rooms for patients with extended-spectrum β-lactamase-producing Enterobacteriaceae in Dutch hospitals: a cluster-randomised, crossover, non-inferiority study. Lancet Infect Dis 2019;19(10):1069–79.

