## Supplementary material for "Regional Variation in the Interpretation of Contact Precautions for Multidrug-resistant Gram-negative bacteria: a cross-sectional survey"

**Table A1** Characteristics of the eleven hospitals from the Southwest region of the Netherlands[1].

| **Characteristics** | **Hospital 1** | **Hospital 2** | **Hospital 3** | **Hospital 4** | **Hospital 5** | **Hospital 6** | **Hospital 7** | **Hospital 8** | **Hospital 9** | **Hospital 10** | **Hospital 11** |
|---|---|---|---|---|---|---|---|---|---|---|---|
| | **University hospital** | **Non-university teaching hospital** | **Non-university teaching hospital** | **Non-university teaching hospital** | **Non-university teaching hospital** | **Non-university teaching hospital** | **Non-university teaching hospital** | **Non-teaching hospital** | **Non-teaching hospital** | **Non-teaching hospital** | **Non-teaching hospital** |
| No. of hospital beds | 1233 (including Sophia children's hospital) | Not available | 564 | 484 | 341 | 332 | Not available | 364 | ≈290 | 194 | 189 |
| No. of single-occupancy rooms | 576 (excluding Sophia children's hospital) | 145 | 80 | 248 | 26 | 84 (including ICU and mother/child centre) | 4 | 49 | 19 | 50 | 33 |
| No. of isolation rooms with anteroom | 54 | 62 | 33 | 66 | 12 | 7 | 0 | 18 | 7 | 10 | 5 |
| Clinical hospital admissions | 30,771 | 32,022 | 21,249 | 24,561 | 27,726 | 13,485 | Not available | 35.475 | ≈10,000 | 16,340 | 9,506 |
| Outpatient clinic visits | 659,317 | 402,758 | 29,5567 | 107,249[2] | 76,729[2] | 266,296[3] | 126,898 | 307,226[3] | ≈50,000 | 42,000 | 165,695 |

*ICU* intensive care unit.

[1] Data are from 2021.

[2] Only number of first outpatient clinic visits available.

[3] Only number of consultations available.

**Table A2** IPC measures recommended by the Dutch, ESCMID and ECDC guidelines.

| | Dutch guidelines | | ESCMID guideline [1] | | ECDC guideline on CPE [2] |
|---|---|---|---|---|---|
| **IPC measures** | Strict isolation [3] | Contact isolation [4] | Contact precautions: endemic MDR-*P. aeruginosa* | Contact precautions: endemic ESBL-E (with the exception of *Escherichia coli*) | Contact precautions |
| **Type of room** | Isolation room[1] | Single-occupancy room, or under exceptional circumstances a multiple-occupancy room provided there is a 1.5 meter space around the patient's bed | Isolation room | Single room | Isolation room with en suite or bathroom facilities designated for use by known carriers, or commodes |
| *Door allowed to be open* | N/A | Yes | N/A | No recommendations | N/A |
| *Patient allowed to leave the room (for other reasons than e.g., operations, radiology)* | No | Yes, under specific conditions | No recommendations | No recommendations | No recommendations |
| **PPE for HCW** | Gloves before entering the room | Gloves during contact with the patient or contact with the patient's direct environment | Gloves before entering the room | Gloves before entering the room | Gloves during direct contact with the patient or their immediate surroundings and/or surfaces. |
| | Long-sleeved gown before entering the room | | Gown before entering the room | Gown before entering the room | Gown/apron during direct contact with the patient or their immediate surroundings and/or surfaces. |
| | Mask (type of mask dependent on the type of | | | | |

| | | | | | |
|---|---|---|---|---|---|
| | microorganism) before entering the room | | | | |
| | Hair cap (only for MRSA) | | | | |
| **Physiotherapy for inpatients** | No recommendations on whether and where physiotherapy can take place, and on recommended PPE use | No recommendations on whether and where physiotherapy can take place, and on recommended PPE use | No recommendations on whether and where physiotherapy can take place, and on recommended PPE use | No recommendations on whether and where physiotherapy can take place, and on recommended PPE use | No recommendations on whether and where physiotherapy can take place, and on recommended PPE use |
| **Visitors of inpatients** | | | | | |
| *PPE* | Mask (type of mask dependent on the type of microorganism) | No specific measures | No recommendations | No recommendations | No recommendations |
| | Gown | | | | |
| *Other* | Visitor is not allowed to visit another patient | No specific measures | No recommendations | No recommendations | No recommendations |
| *Rooming-in* | No recommendations | No recommendations | No recommendations | No recommendations | No recommendations |
| **Cleaning and disinfection** | Daily cleaning of the patient room and anteroom and terminal cleaning and disinfection before admission of a new patient | Daily cleaning of the patient room and terminal cleaning and disinfection before admission of a new patient | Implement regular environmental cleaning procedures, which include detergents or disinfectants, depending on local practice to reduce the transmission rate. Ensure cleaning of patient | Implement regular environmental cleaning procedures, which include detergents or disinfectants, depending on local practice to reduce the transmission rate. Ensure cleaning of patient | Enhanced cleaning should be performed, especially for areas in close proximity to CRE carriers. Terminal disinfection of rooms should be performed upon transfer or discharge of patients. |

| | | | |
|---|---|---|---|
| | | care equipment and the environment. | care equipment and the environment. |

*ECDC* European Centre for Disease Prevention and Control, *ESBL-E* extended-spectrum β-lactamase-producing Enterobacterales, *ESCMID* European Society of Clinical Microbiology and Infectious Diseases, *HCW* healthcare worker, *IPC* infection prevention and control, *MDR-P.* aeruginosa multidrug-resistant *Pseudomonas aeruginosa, MRSA* meticillin-resistant *Staphylococcus aureus*, N*/A* not applicable, *PPE* personal protective equipment.

[1] An isolation room is defined as a single-occupancy room with anteroom and pressure differences [5].

**References**

[1] Tacconelli E, Cataldo MA, Dancer SJ, De Angelis G, Falcone M, Frank U, et al. ESCMID guidelines for the management of the infection control measures to reduce transmission of multidrug-resistant Gram-negative bacteria in hospitalized patients. Clin Microbiol Infect 2014;20 Suppl 1:1-55.
[2] Magiorakos AP, Burns K, Rodríguez Baño J, Borg M, Daikos G, Dumpis U, et al. Infection prevention and control measures and tools for the prevention of entry of carbapenem-resistant Enterobacteriaceae into healthcare settings: guidance from the European Centre for Disease Prevention and Control. Antimicrob Resist Infect Control 2017;6:113.
[3] Dutch Working Party on Infection Prevention (WIP). Strikte isolatie. 2006.
[4] Dutch Working Party on Infection Prevention (WIP). Contactisolatie. 2006.
[5] Dutch Working Party on Infection Prevention (WIP). Bouw- en inrichtingseisen isolatie-afdeling Ventilatie isolatiekamers. 2004.

**Table A3** IPC measures for visitors of inpatients (n=11 hospitals).

| Transmission-based precautions | CPE | CPPA | ESBL-E |
|---|---|---|---|
| **IPC measures for visitor(s)**[1] | 11 (100) | 11 (100) | 10 (90.9) |
| Hand disinfection | 11 (100) | 11 (100) | 10 (100) |
| Disposable gown and non-sterile gloves | 1 (9.1) | 1 (9.1) | 1 (10.0) |
| Disposable gown | 1 (9.1) | 1 (9.1) | 0 (0.0) |
| Surgical mask type IIR, disposable gown, non-sterile gloves, and hair cap | 1 (9.1) | 1 (9.1) | 0 (0.0) |
| **Temporary room leave by visitor(s) for a toilet visit is allowed** | | | |
| Yes | 4 (36.4) | 4 (36.4) | 5 (45.5) |
| No | 6 (54.5) | 6 (54.5) | 4 (36.4) |
| Not applicable[2] | 1 (9.1) | 1 (9.1) | 2 (18.2) |
| **Temporary room leave by visitor(s) for use of the coffee area is allowed** | | | |
| Yes | 4 (36.4) | 4 (36.4) | 6 (54.5) |
| No | 6 (54.5) | 6 (54.5) | 3 (27.3) |
| Not applicable[2] | 1 (9.1) | 1 (9.1) | 2 (18.2) |
| **Rooming-in is allowed** | | | |
| Yes | 10 (90.9) | 10 (90.9) | 9 (81.8) |
| No | 1 (9.1) | 1 (9.1) | 1 (9.1) |
| Not applicable | 0 (0.0) | 0 (0.0) | 1 (9.1) |
| **Temporary leave of rooming-in visitor from patient room is allowed** | | | |
| Yes | 5 (45.5) | 5 (45.5) | 4 (36.4) |
| No | 5 (45.5) | 5 (45.5) | 5 (45.5) |
| Not applicable | 1 (9.1) | 1 (9.1) | 2 (18.2) |

Data are N (%). *CPPA* carbapenemase-producing *Pseudomonas aeruginosa, CPE* carbapenemase-producing Enterobacterales, *ESBL-E* extended-spectrum β-lactamase-producing Enterobacterales*, IPC* infection prevention and control, *N/A* not applicable.

[1] Multiple answers per hospital possible.

[2] One hospital has no specific policy on this topic in place.

**Table A4** Cleaning and disinfection after discharge of the patient (n=11 hospitals).

| | CPE | CPPA | ESBL-E |
|---|---|---|---|
| **Cleaning product for final cleaning**[1,2] | | | |
| Reusable microfiber cloth | 2 (18.2) | 2 (18.2) | 2 (18.2) |
| Reusable mop | 2 (18.2) | 2 (18.2) | 2 (18.2) |
| Disposable (microfiber) cloth | 5 (45.5) | 5 (45.5) | 5 (45.5) |
| Disposable mop | 5 (45.5) | 5 (45.5) | 5 (45.5) |
| No separate cleaning[3] | 3 (27.3) | 3 (27.3) | 2 (18.2) |
| Not applicable | 0 (0.0) | 0 (0.0) | 1 (9.1) |
| **Method or product for final disinfection**[1,2] | | | |
| Chlorine 250 ppm | 5 (45.5) | 5 (45.5) | 5 (45.5) |
| Chlorine 1000 ppm | 2 (18.2) | 2 (18.2) | 1 (9.1) |
| Alcohol 70% | 6 (54.5) | 6 (54.5) | 4 (36.4) |
| 2-in-1 cleaning and disinfection product | 3 (27.3) | 3 (27.3) | 2 (18.2) |
| Hydrogen peroxide wipes | 1 (9.1) | 1 (9.1) | 1 (9.1) |
| No disinfection | 0 (0.0) | 0 (0.0) | 1 (9.1) |
| Not applicable | 0 (0.0) | 0 (0.0) | 1 (9.1) |
| **Changing separation curtains after isolation** | | | |
| Yes | 9 (81.8) | 9 (81.8) | 7 (63.6) |
| No | 2 (18.2)[4] | 2 (18.2)[4] | 3 (27.3)[4] |
| Not applicable | 0 (0.0) | 0 (0.0) | 1 (9.1) |
| **Changing window curtains after isolation** | | | |
| Yes | 1 (9.1) | 1 (9.1) | 0 (0.0) |
| No | 10 (90.9)[4] | 10 (90.9)[4] | 10 (90.9)[4] |
| Not applicable | 0 (0.0) | 0 (0.0) | 1 (9.1) |

Data are N (%). *CPPA* carbapenemase-producing *Pseudomonas aeruginosa, CPE* carbapenemase-producing Enterobacterales, *ESBL-E* extended-spectrum β-lactamase-producing Enterobacterales.

[1] Multiple answers per hospital possible.

[2] Missing data from one hospital.

[3] No separate cleaning due to the use of a 2-in-1 cleaning and disinfection product.

[4] One hospital does not generally change the separation or window curtains after cessation of isolation measures, but only in case of visible contamination or if the patient provides a reason to do so (e.g., demented patient).
